# Simulating the progression of the COVID-19 disease in Cameroon using SIR models

**DOI:** 10.1101/2020.05.18.20105551

**Authors:** Ulrich Nguemdjo, Freeman Meno, Audric Dongfack, Bruno Ventelou

## Abstract

This paper analyses the evolution of COVID-19 disease in Cameroon over the period March 6—April 2020 using SIR model. Specifically, 1) we evaluate the basic reproduction number of the virus. 2) Determine the peak of the infection and the spread-out period of the disease. 3) Simulate the interventions of public health authorities. Data used in this study is obtained from the Ministry of health of Cameroon. The results suggest that over the period, the reproduction number of the covid-19 in Cameroon is about 1.5 and the peak of the infection could occur at the end of May 2020 with about 7.7% of the population infected. Besides, implementation of efficient public health policies could help flattens the epidemic curve.

## 1. Introduction

The new coronavirus (COVID-19) started in Wuhan, China last November showing pneumonia-like symptoms. The first cases examined in China indicated that it was a new respiratory disease. However, The World Health Organization (WHO) took to delivering the most recent discoveries about this virus from January 7, 2020. By the end of January 2020, although the virus had already been spread in few countries, WHO alleged the World announcing an international sanitary crisis.

By February 14, 2020, the WHO (2020) reported that the first confirmed case on the African continent was registered in Egypt whereas the ministry of health of Cameroon announced their first confirmed on March 6, 2020. Following this announcement regarding the first confirmed case the widespread of the diseases have been gradually increasing within the population. Based on the current situation there is need to study the evolution of this virus and the efficiency of the measures adopted by local authorities to curb the spread of Covid-19 diseases in Cameroun. The rest of the study is organized as follows. We present the data in section 2. Section 3 outlines the model. Results are presented in section 4. Finally, section 5 concludes.

## 2. Data

To explore the evolution of the coronavirus disease in Cameroon, the present paper uses data collected from the Cameroonian ministry of health. According to the government, the first confirmed case of the COVID-19 was detected on March 6, 2020, in the political capital, Yaoundé. The government decided to publish daily report regarding the evolution of the disease including: the number of confirmed cases of COVID-19, the number of death due to the virus and the number of recoveries via public declaration. These declarations ended on April 10, 2020, that is why our database goes from March 6 to April 10.

Figure 1 shows an overview of the evolution of coronavirus disease in Cameroon during our period of study. It shows an exponential increase in the number of confirmed cases. The total number of confirmed cases increase from 10 on March 17 to 820 on April 10, that is multiplying the total number of cases by 82 in less than a month. Moreover, the figure shows a slight increase in the total number of deaths due to COVID-19 in the country, from 1 on March 25 (the first case) to 12 on April 10. On the last day of our period of observation, the figure shows that the total number of recoveries from COVID-19 in Cameroon is about 98 which is around 8 times more than the total number of deaths observed on the same date.

**Figure 1:**
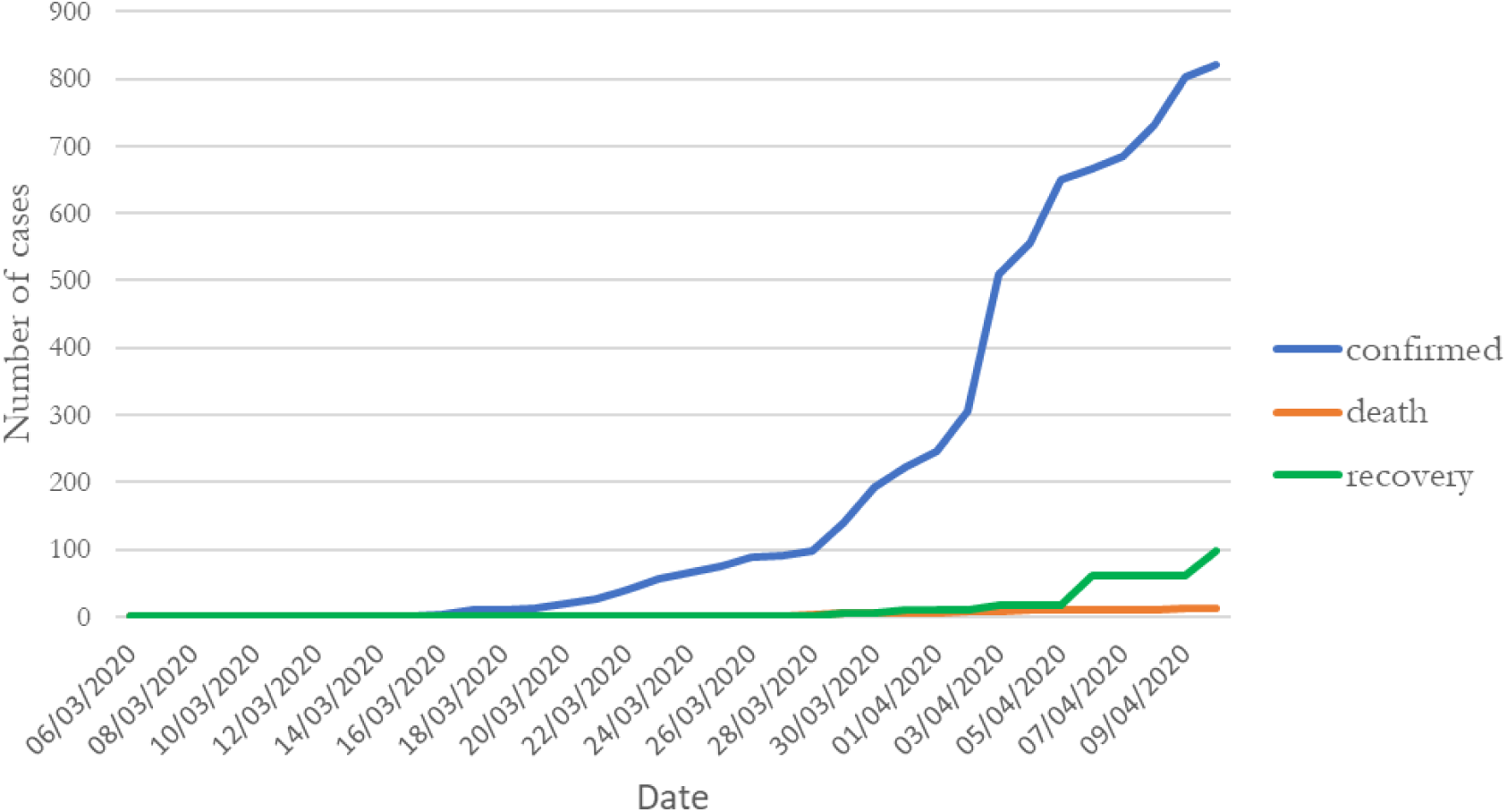
Number of observed cases of COVID-19 in Cameroon

## 3. The Model

To explore the evolution of the coronavirus disease in Cameroon, this paper uses a simple SIR (Susceptible - Infectious - Recovered) model known as a general stochastic epidemic model (Kendrick and Kermarck 1927; Jones 2007). This model is particularly suitable while dealing with a large population. The initial model considers that individuals are at first Susceptible. If they get infected by the virus, they become immediately Infectious) and they remain infectious until they Recover assuming immunity during the rest of the outbreak. In our paper, the last group was modified to “Removed” (Giraldo and Palacio, 2008). Thus, the individual is Infectious until he is Removed either by being placed in quarantine, being hospitalized or by recovering (and not susceptible anymore^5^) or dying from the virus. We also assume that the population in our study is closed and that individuals mix uniformly in the community. Following Britton and Giardina (2016) we also assume that all individuals are equally susceptible to the disease and equally infectious if they get infected.

### 3.1. The SIR Model

Supposed a closed population of size N, an individual who gets infected by the coronavirus becomes immediately infectious and remains so for an exponentially distributed time with rate parameter γ (the removal rate). Thus, *γ*^−1^refers to the average number of infectious days that an infected individual has to transmit the virus before being removed from the Infectious group (placed in quarantine, hospitalized or recovered or died). During the infectious period, an individual has “close contact” with the rest of the population in time at rate β. The parameter β = τ × μ known as the effective contact rate can be written as the product of two other parameters: the average number of exposure per unit of time (τ) and the likelihood of infection at each occasion of exposure (μ) (Jones, 2007).

*S(t), I(t)* and *R(t)* respectively represents the number of susceptible, infectious, and removed individuals in the population at time *t*. Assuming that we have a closed population, the total population at time period t is given as follows:

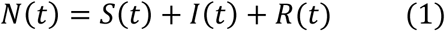

Let’s assume that at the beginning of the epidemic *(S(0), I(0), R(0)) = (N-1, 1, 0)* meaning that there is initially one infectious individual in the population and no removal. All other things being equal, the number of susceptible individuals decreases symmetrically by:

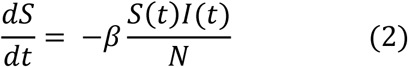

Also, the variation in the number of people infected according to this measure is given by:

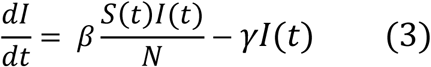

The result is the following dynamic system, reflecting the generalized SIR Model:

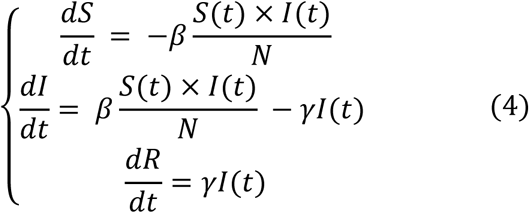

The simplicity of this dynamic system gives us rapid information on the rate of spread of the epidemic. Indeed, an epidemic occurs if the number of infected individuals increases continuously, i.e., 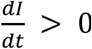

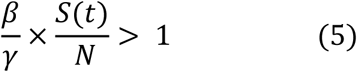

As highlighted by Jones (2007), at the outset of an epidemic, nearly everyone is susceptive. Thus, 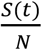 can be approximated to 1 and the above equation can be written as:

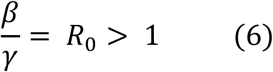

### 3.2. The Parameter R_0_

The parameter R_0_ is called the basic reproduction rate. It is the expected number of secondary cases produced by a single infectious individual during the infectious period in a completely susceptible population (Jones, 2007). Regarding the value of the parameter, the severity of the epidemic can be summarized into two cases (Britton and Giardina, 2016):

- R_0_ > 1: The Supercritical case. The epidemic increases exponentially: one infected individual infects more than one individual on average.
- R_0_ ≤ 1: Critical case. No epidemic: the disease will surely die out without affecting a large share of the population

Note that an R_0_ > 1 does not always guarantee an epidemic in the population (Choisy et al. 2007; Bartlett, 1960)

## 4. The results

### 4.1. Estimating the parameters of the model

To simulate the progression of the coronavirus disease in Cameroon, the next step consists in determining the parameters *β* and *γ* that best describe the current evolution of the virus in Cameroon^6^ (presented in section 2). This is done using a Nelder-Mead and Maximum likelihood optimization algorithm. The process implies finding *β* and *γ* in such a way that when these parameters are substituted in the equations described above, the difference between the data obtained and that recorded on the field is minimized. Computing *β* and *γ* using data recorded from March 6 to April 10 and a population of size *N = 25,216,237*. The results are summarized in Table 1. Simulations conducted with the above parameters yields the results shown in Figure 2.

**Table 1:** Estimates parameters^7^

|  | Original | Bias | Std. error | Bootstrap normal CI* |  |
| --- | --- | --- | --- | --- | --- |
|  |  |  |  | Inf | Sup |
| $\beta$ | 0.615 | 7.65e-06 | 0.003 | 0.610 | 0.619 |
| $\gamma$ | 0.393 | -3.69e-05 | 0.003 | 0.388 | 0.398 |
| $R_0$ | 1.567 | 0.000 | 0.016 | 1.536 | 1.597 |
| Maximum Infected | 2,015,200 | 757.6864 | 76,463.73 | 1,864,576 | 2,164,309 |
| Number of Days to reach the peak | 81,06 | -0.022 | 1.660 | 77.81 | 84.32 |

\*CI = Confidence Interval
<sup>6</sup> The simulations are carried out mindless of the measures adopted by the Cameroonian local government to obstruct the way to the spread of the virus
<sup>7</sup> The distributions of the bootstrap realizations are presented in Appendix (Figures 4 to 9)

**Figure 2:**
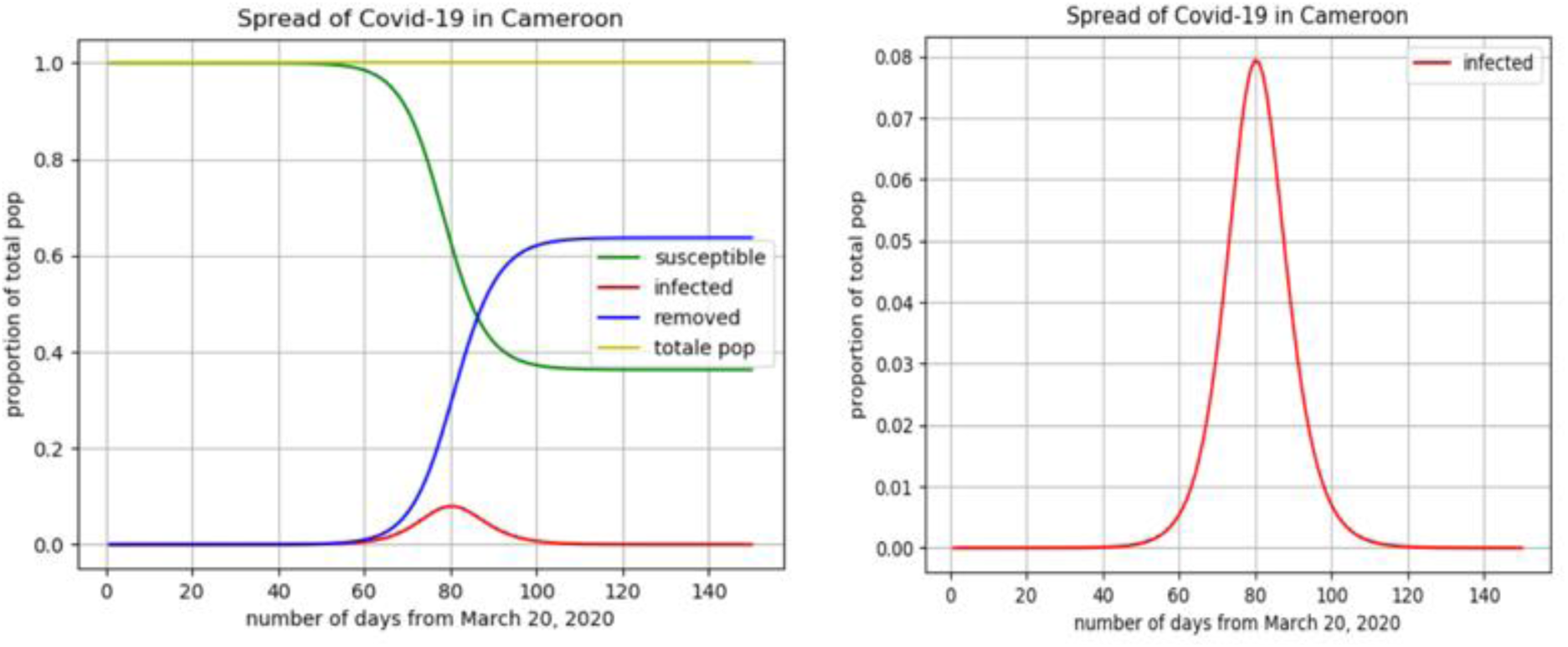
evolution of the coronavirus disease in Cameroon

All else unchanged, the evolution presented in Figure 2 should be a plausible scenario if nothing is done indeed to reduce the spread of the virus. More precisely, the figure reveals that, if the situation remains as during the observed period (from Marche 6 to April 10), about 7.7% of the Cameroonian population might have ended up being infected, which is close to *2,015,200* individuals. In this case, the peak of the infection will occur between day number 78 and day number 85 starting from March 6, which is between 25th and 29th May. Additionally, assuming a rate of 15.8% of serious complications of the disease (Petrili et al. 2020) approximately *318,402* Cameroonians might find themselves hospitalized in critical conditions. While considering a mortality rate of 3.4% (the overall mortality rate of COVID-19 announced by the WHO on March 3, 2020^8^) the expected number of deaths due to coronavirus could be close to *68,517*. Though these results might not be accurate, the approach gives a simple and quick means to figure out the evolution of the spread of the virus and paves the way for a better approach. Our model also suggests that the basic reproduction rate (R_0_) of the COVID-19 in Cameroon up to April 10 is about 1.567 persons meaning that on average, an infectious individual infects 1.567 susceptible individuals during his infectious period.

The question now is how can we flatten the infectious curve? What has been recommended since the beginning of this epidemic is to apply public health measures.

### 4.2. Flattening the epidemic curve with public health interventions

Until an efficient medical treatment or vaccine for COVID-19 is available, prevention and control strategies to reduce or stop the transmission of the disease only rely on measures adopted by public health officials. In this section, we are modeling the effect of different public health interventions on the spread of the coronavirus (COVID-19) disease in Cameroon.

#### Physical distancing

Physical or social distancing-keeping space between yourself and other people outside your home- plays a major role in public health interventions. Its objective is to reduce the probability of contact between infected individuals and susceptible ones, to minimize the transmission of the disease. In practice, physical distancing^9^ can be implemented by adopting the following habits:

- Stay at least 1 meter (3 feet) from others
- Do not gather in groups
- Stay out of crowded places
- Avoid mass gathering

#### Hygiene measures

In addition to physical distancing, public health interventions also incorporate hygiene measures to reduce transmission of the coronavirus between individuals, from individuals to surfaces and from surfaces to other individuals. These hygiene measures include:

- Wash hands frequently
- Avoid touching eyes, nose, and mouth
- Practice respiratory hygiene: cover the mouth and the nose with the bent elbow or tissue when coughing or sneezing
- Wearing surgical masks
- Cleaning or disinfection of fomites

#### Simulating public health interventions

The two main public health interventions mentioned above can be implemented in our model by adjusting two parameters (Churches, 2020). More precisely, to simulate an increase in social or physical distancing, we can reduce the average number of exposures per day (τ), whereas to take into consideration the hygiene measures, we can reduce the probability of infection at each occasion of exposure (μ) of the parameter β of the SIR model (see section 3.1).

According to some studies on the COVID-19, the probability of infection at each exposure varies from 1% to 5% (Tang et al., 2020). In this paper, 5% is selected as the initial infection probability. As our parameter β = 0.6, we can estimate the initial average number of exposures per day to 12 exposures. Therefore, the goal of the public health interventions in Cameroon will be to reduce progressively the number of exposure (we will simulate using 12, 6, and 2 exposures per day) and the probability of infection (we will simulate using 5%, 2.5%, and 1%)^10^. Figure 3 gives the results of the simulations.

**Figure 3:**
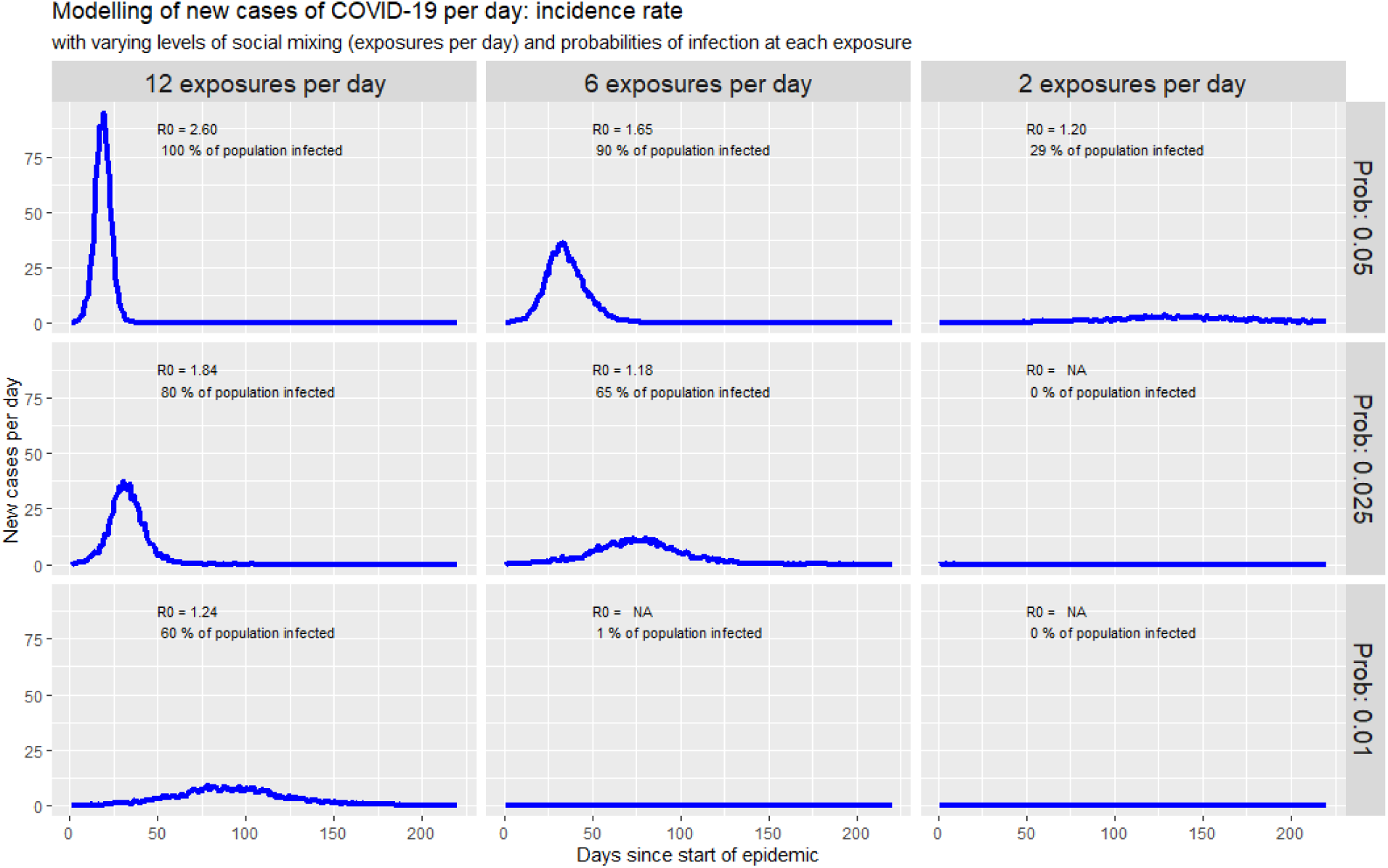
Simulating new cases of COVID-19 with public health interventions

As we can notice, a decrease in the number of exposures per day (from the left to the right on the figure) and a decrease in the likelihood of infection (from the top to the bottom of the figure) is associated with a flattening in the coronavirus epidemic curve. Besides, we can observe a decrease in the percentage of the population infected by the virus and the disease lasts for a shorter period.

## 5. Conclusion

The present paper aims to analyze the evolution of the coronavirus disease in Cameroon. The study uses data collected by the Cameroonian health ministry between March 6 (date of the first confirmed case in the country) to April 10, 2020.

Descriptive statistics show an exponential increase in the total number of infectious individuals. This gives a first idea of how the virus is spreading out in Cameroun. Starting from this natural evolution of the epidemic in the first days of propagation in the country, a SIR model was calibrated on the observed data. The results of the calibration show that, if actions undertaken by Cameroonians to fight against the coronavirus do not improve, the peak of the infection will occur at the end of May 2020 with about 7.7% of the Cameroonian population infected. Using the World Health Organization (WHO) mortality rate associated with the COVID-19, the expected number of deaths due to the virus in Cameroun could be close to *70,000*.

However, by intensifying public health interventions in the simulation, the epidemic curve flattens. These results seem to underline the importance of appropriate communication campaigns from the government and of the compliance, by the population, with the public health measures recommended by the World Health Organization (WHO) in order to limit and stop the spread of the coronavirus disease at least while waiting for a possible preventive and/or curative treatments to be found.

Even though the paper has some limitations, as the closed population assumptions, the homogeneous mixture of the population (particularly across the geography of Cameroun), that may be difficult to observe in the real life situations, our model is particularly suitable while dealing with a large population, as a population observed at a country level (Jones, 2007). In all cases, knowing the paucity of the literature available for African countries, this paper enriches the knowledge by providing some quantitative evidence in support of the Cameroonian government actions that attempt to fight against the coronavirus. As an extension of this paper, studies with more sophisticated assumptions on the contact-matrix (between susceptible) for the epidemiology models could be carried out as well as simulations showing the economic impact of the coronavirus epidemic on the Cameroonian population.

## Data Availability

Data used for the study are daily information collected through the report of the Cameroonian health minister. They can also be found on the link below.

https://louisdecharson.github.io/covid-19

## Appendix

**Figure 4:**
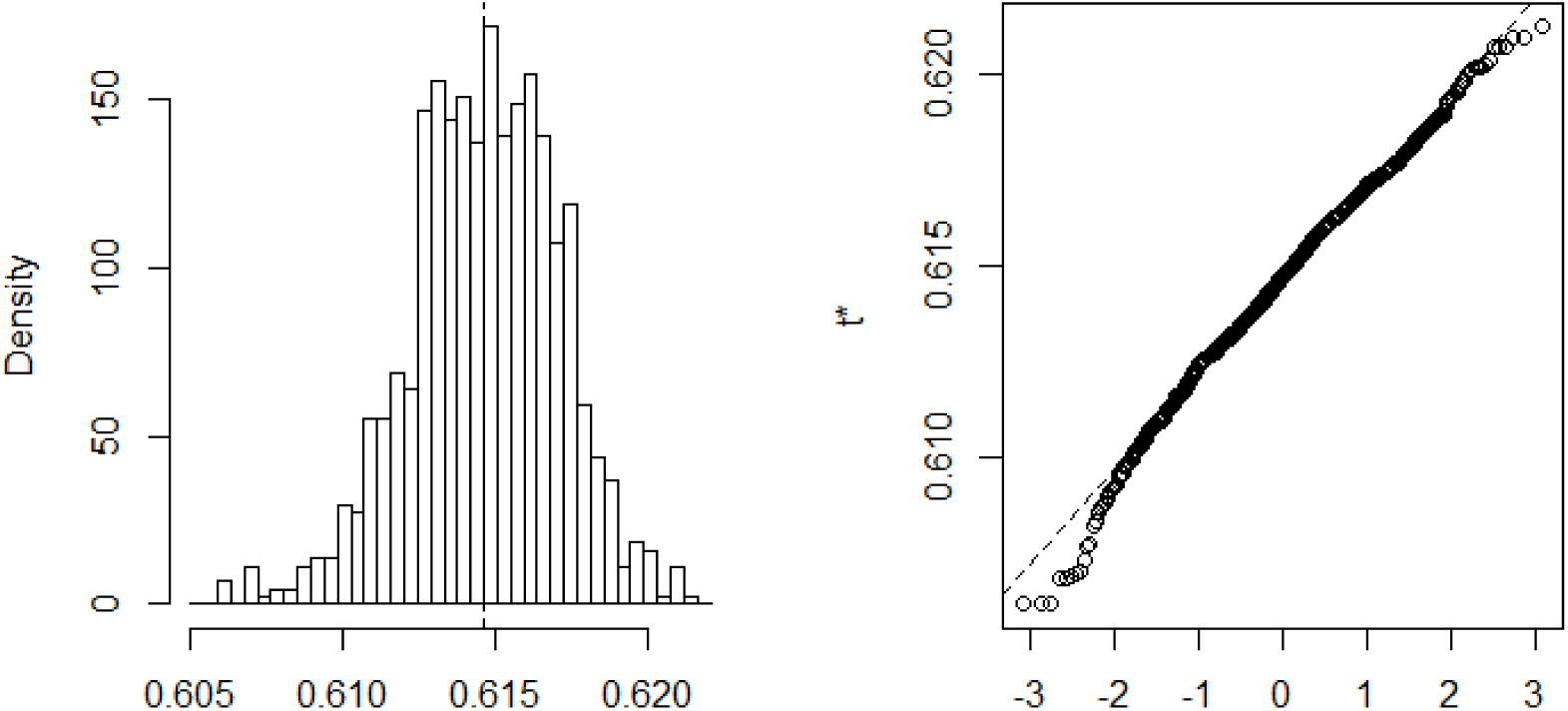
Bootstrap distribution of the β

**Figure 5:**
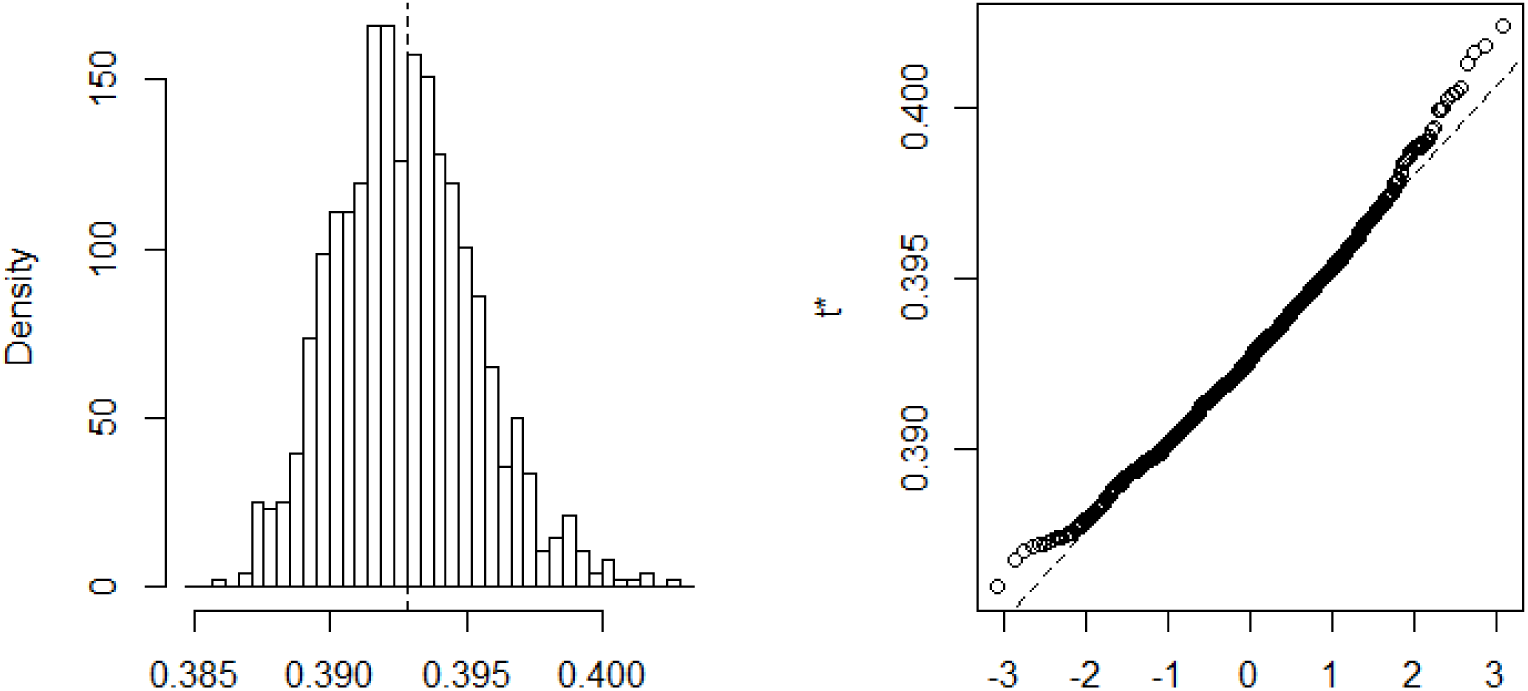
Bootstrap distribution of the γ

**Figure 6:**
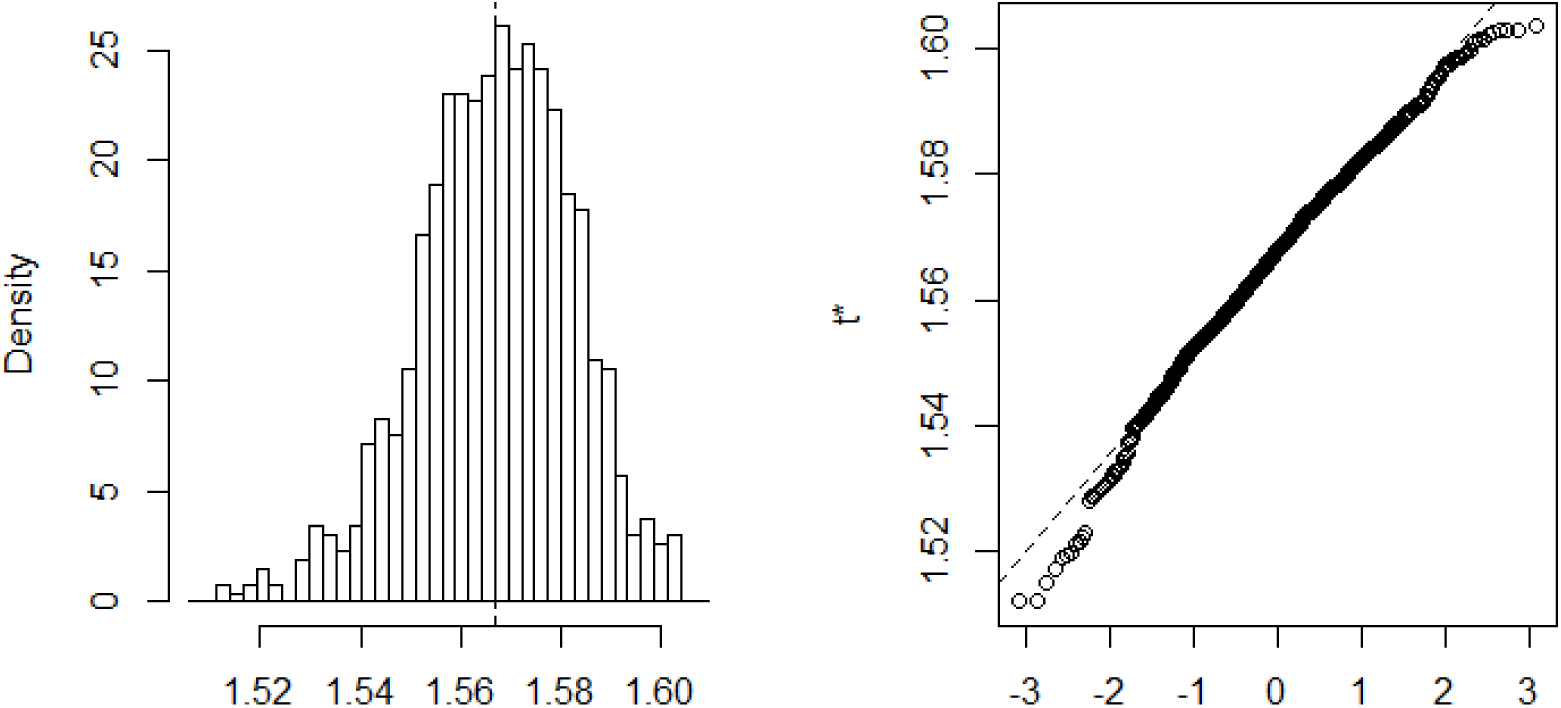
Bootstrap distribution of the *R*_0_

**Figure 8:**
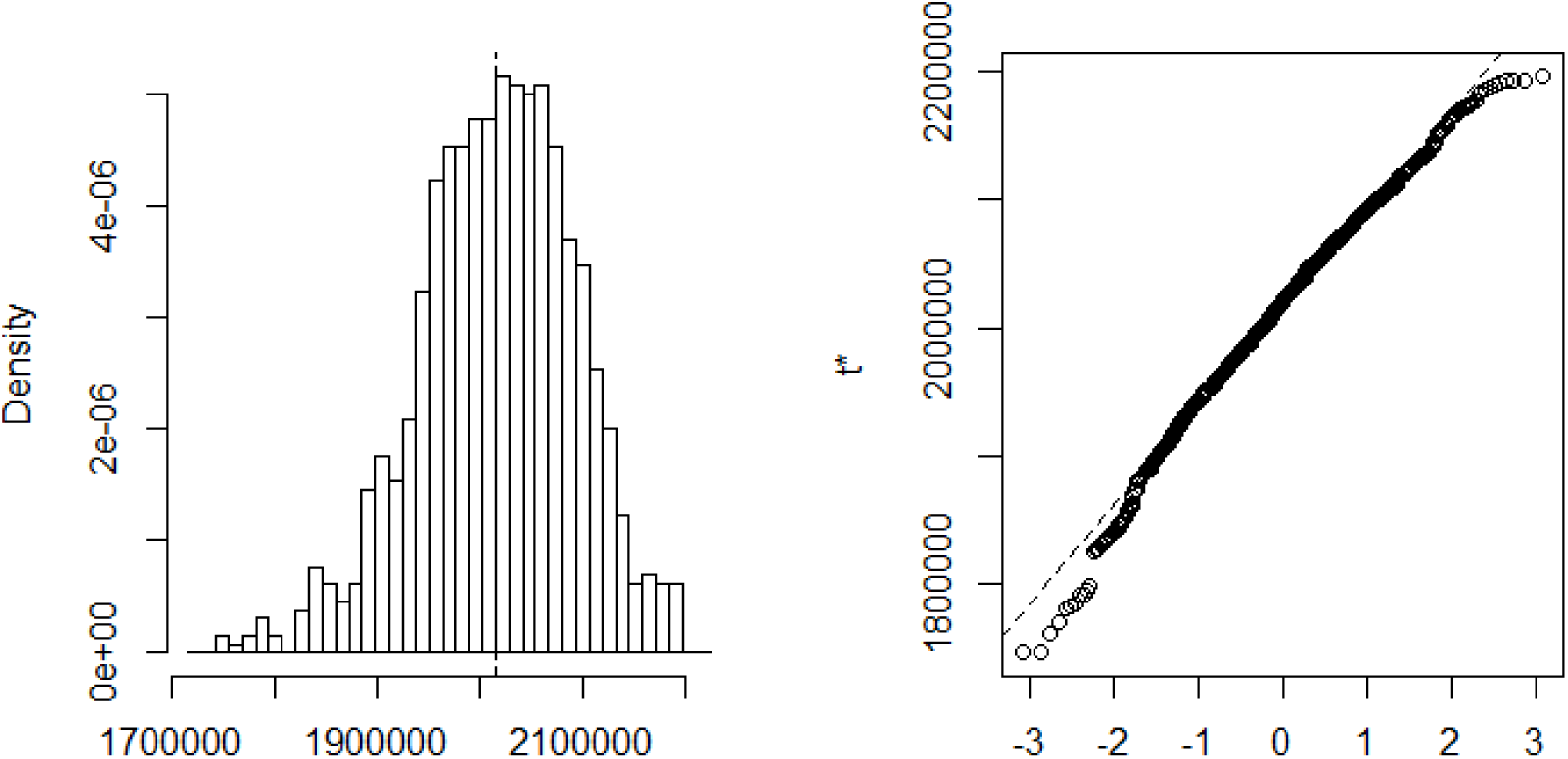
Bootstrap distribution of the infected

**Figure 9:**
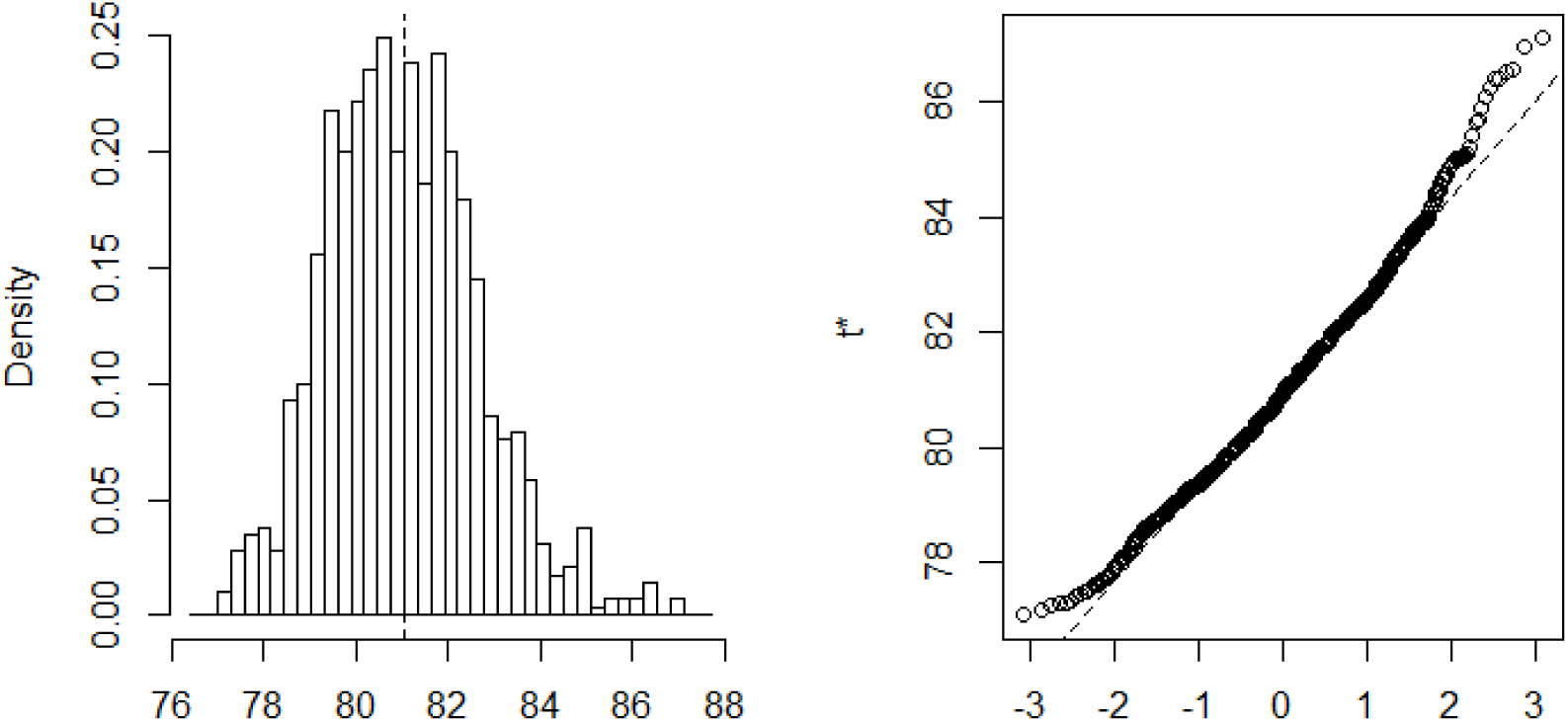
Bootstrap distribution of the number of days

## Footnotes

5 Up till now (April 10, 2020), it is not sure that, in the case of COVID-19, the recovered will be immunized for a very long period. In this paper, we assume that recovery confers immunity during the rest of the outbreak.

6 The simulations are carried out mindless of the measures adopted by the Cameroonian local government to obstruct the way to the spread of the virus

7 The distributions of the bootstrap realizations are presented in Appendix (Figures 4 to 9)

8 https://www.who.int/dg/speeches/detail/who-director-general-s-opening-remarks-at-the-media-briefing-on-covid-19---3-march-2020

9 https://www.who.int/emergencies/diseases/novel-coronavirus-2019/advice-for-public

10 We used the R *EpiModel* package to simulate public interventions with a population of size N = 1,000 for computational matters (Jenness et al. 2018).

